# A personalized antibody score for predicting individual COVID-19 vaccine-elicited antibody levels from basic demographic and health information

**DOI:** 10.1101/2022.07.05.22277283

**Authors:** Naotoshi Nakamura, Hyeongki Park, Kwang Su Kim, Yoshitaka Sato, Yong Dam Jeong, Shoya Iwanami, Yasuhisa Fujita, Tianchen Zhao, Yuta Tani, Yoshitaka Nishikawa, Chika Yamamoto, Yurie Kobashi, Takeshi Kawamura, Akira Sugiyama, Aya Nakayama, Yudai Kaneko, Kazuyuki Aihara, Shingo Iwami, Masaharu Tsubokura

## Abstract

Antibody titers wane after two-dose COVID-19 vaccinations, but individual variation in vaccine-elicited antibody dynamics remains to be explored. Here, we created a personalized antibody score that enables individuals to infer their antibody status by use of a simple calculation. We recently developed a mathematical model of B cell differentiation to accurately interpolate the longitudinal data from a community-based cohort in Fukushima, Japan, which consists of 2,159 individuals who underwent serum sampling two or three times after a two-dose vaccination with either BNT162b2 or mRNA-1273. Using the individually reconstructed time course of the vaccine-elicited antibody response, we first elucidated individual background factors that contributed to the main features of antibody dynamics, i.e., the peak, the duration, and the area under the curve. We found that increasing age was a negative factor and a longer interval between the two doses was a positive factor for individual antibody level. We also found that the presence of underlying disease and the use of medication affected antibody levels negatively, whereas the presence of adverse reactions upon vaccination affected antibody levels positively. We then applied to these factors a recently proposed computational method to optimally fit clinical scores, which resulted in an integer-based score that can be used to evaluate the antibody status of individuals from their basic demographic and health information. This score can be easily calculated by individuals themselves or by medical practitioners. There is a potential usefulness of this score for identifying vulnerable populations and encouraging them to get booster vaccinations.

**Significance statement:** Different individuals show different antibody titers even after the same COVID-19 vaccinations, making some individuals more prone to breakthrough infections than others. Such variability remains to be clarified. Here we used mathematical modeling to reconstruct individual post-vaccination antibody dynamics from a cohort of 2,159 individuals in Fukushima, Japan. Machine learning identified several positive and negative factors affecting individual antibody titers. Positive factors included adverse reactions after vaccinations and a longer interval between two vaccinations. Negative factors included age, underlying medical conditions, and medications. We combined these factors and developed an “antibody score” to estimate individual antibody dynamics from basic demographic and health information. This score can help to guide individual decision-making about taking further precautions against COVID-19.

## Text

The ongoing COVID-19 pandemic has caused more than 500 million cases and 6 million confirmed deaths worldwide. The current COVID-19 vaccines, which became available in late 2020 to early 2021, have helped vaccine recipients acquire immunity against SARS-CoV-2 and reduce their likelihood of infection and hospitalization (1, 2).

In the case of two-dose mRNA-based vaccines such as BNT162b2 and mRNA-1273, antibody titers, on average, reach a peak around two weeks after the second vaccination and decline thereafter. However, explanation of the individual variability in vaccine-elicited antibody dynamics has remained elusive. Past studies addressing dynamics focused on the average of a group of individuals (i.e., population-level dynamics) having specific demographic characteristics such as age or sex (3-8) and thus have not led to personalized advice for individuals. Some studies included only health care workers and did not cover the whole spectrum of the general population, especially older adults and those with underlying medical conditions (4, 9-12). Most of the other studies targeted at the general population lumped antibody measurements together with different elapsed times since vaccination (12-17), making precise determination and comparison of individual dynamics difficult.

Here, we used a mathematical model (developed in (18)) to describe the process of differentiation from naïve B cells to plasma cells to accurately reconstruct individual vaccine-elicited antibody dynamics in the Fukushima vaccination cohort (a community-based cohort in Fukushima, Japan). The model parameters describe highly variable individual-level antibody responses, allowing us to partially predict variation in vaccine response on the basis of personal information including age, adverse reactions, comorbidities, and medication use. Furthermore, we devised a useful personalized antibody score that allows individuals to predict their antibody status from their personal information. The score can be used by medical practitioners to encourage individuals with low predicted antibody levels to get booster vaccinations.

## Results

### Deriving measures of peak, duration, and area under the curve of vaccine-elicited antibody dynamics

We fully reconstructed the dynamics of IgG(S) titers after the first vaccination for 2,159 individuals in the Fukushima vaccination cohort in **Supplementary Fig 1C** (see **Methods** in detail) and extracted the “features” described in **Fig 1A** for each individual: the peak, duration, and area under the curve (AUC) of the reconstructed antibody dynamics. To quantify these features, we here assumed *A*_TH = 100_ and determined *t*_*s*_ and *t*_*e*_ corresponding to the time for the antibody titer to be greater than and smaller than *A*_TH_, respectively. Therefore, the duration and AUC of the antibody titer are formulated by *t*_*e*_ − *t*_*s*_ and 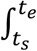*A*(*s*)*ds*, respectively. In addition, defining *t*_*p*_ to be the time for the antibody titer to reach its peak, the peak titer is *A*(*t*_*p*_). In **Fig 1B**, we summarized distributions of the AUC, duration, and peak for 2,159 participants. We then compared these features among the participants. We used the logarithm (log_10_) of the peak and of the AUC because these measures had a long-tailed distribution spanning at least two orders of magnitude (**Supplementary Fig 2A**). The Pearson correlation matrix (**Fig 1C**) showed that the AUC was highly correlated with the duration and the peak, meaning that the three features are similar. Note that a similar trend was obtained under a different *A*_TH_ (data not shown). These features allowed us to quantitatively compare vaccine-elicited antibody dynamics among the participants (see next section).

**Figure 1.**
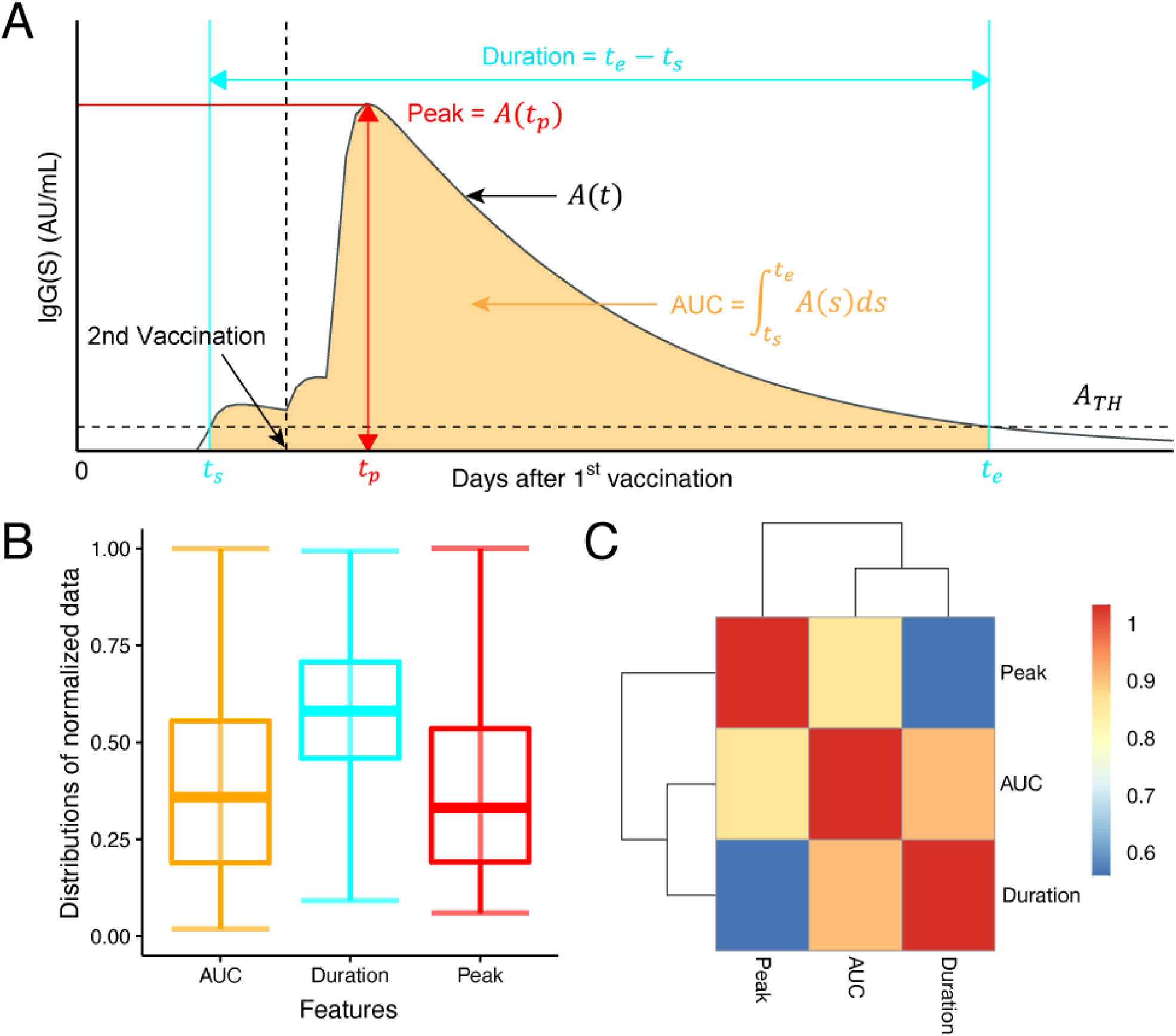
Quantifying vaccine-elicited antibody dynamics: **(A)** Vaccine-elicited antibody response after the first vaccination (i.e., *t* = 0) is described with the following “features”: the peak (*A*(*t*_*p*_)), duration (*t*_*e*_− *t*_*s*_), and AUC 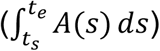 of the antibody titers. The vertical and horizontal dashed lines correspond to the date of the second vaccination and the arbitrary threshold (*A*_TH_) for calculating the duration and AUC, respectively. **(B)** Distributions of the extracted features from the reconstructed antibody dynamics (i.e., the peak, duration, and AUC) for 2,407 participants are plotted. The dataset for each distribution was normalized by the value corresponding to the 95th percentile of data values, and data whose values were larger than this value were removed to improve the visibility of the figure. **(C)** Heatmap plot showing Pearson correlation matrix describing three features (log_10_ of peak, log_10_ of AUC, duration) of antibody dynamics.

### Characterizing vaccine-elicited antibody dynamics

To see how individual background factors contributed to the three features, we trained a random forest regressor to predict the features from basic demographic information for the participants, including underlying medical conditions, adverse reactions to vaccinations, and medications, as described in **Supplementary Table 1**. The out-of-bag (OOB) R squared values obtained were 15.9%, 27.9%, and 23.7% for the peak, AUC, and duration, respectively, suggesting that the features can be partially explained by the background information available. We visualized significant factors as a Chord diagram, in which the size of the arcs connecting individual factors to each of the three features indicates their importance in predicting the feature (**Fig 2A**).

**Figure 2.**
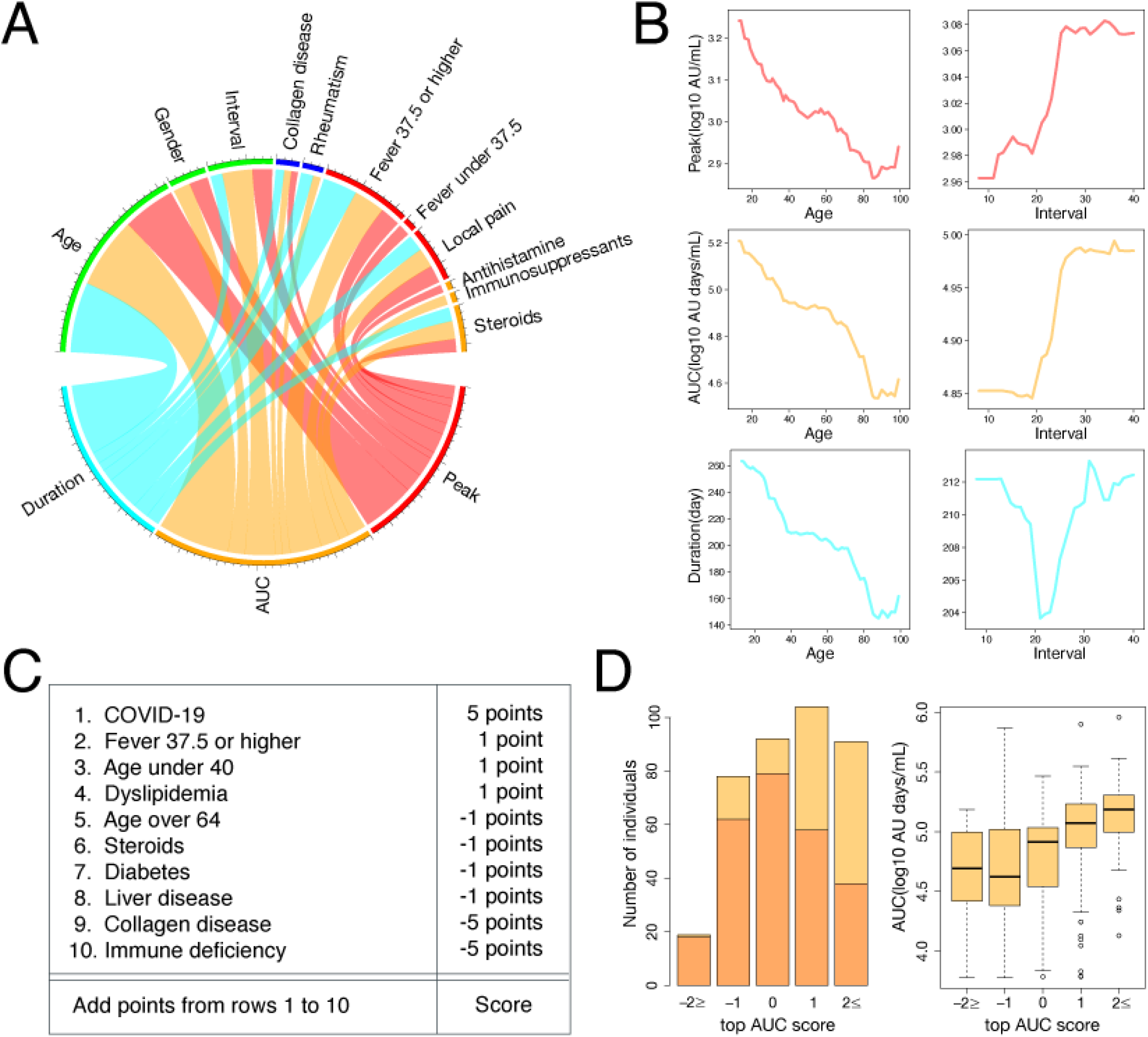
Characterizing and scoring vaccine-elicited antibody dynamics: **(A)** Chord diagram representing the most predictive factors of three “features” of antibody titers (i.e., peak, duration, and AUC) is shown. The size of the arc from a group to a feature is proportional to its importance as measured by Mean Decrease in Accuracy. Features with *p* < 0.05 are displayed. **(B)** Partial dependence plots showing the dependence of the three features on age or the interval between two vaccinations are shown. **(C)** Metric for calculating the “top AUC score,” i.e., a score to identify individuals with AUC in the top third of the population. **(D)** Left: Distribution of the top AUC scores in the test dataset: 19, 78, 92, 104, and 91 individuals had scores of -2 or less, -1, 0, 1, or 2 or more, respectively. Those in the top third of the test dataset are shown in yellow, and those not in the top third are shown in orange. The ratio of individuals with AUC in the top third of the test dataset increased as the top AUC score increased. Right: the average AUC tended to increase as the top AUC score increased.

We used partial dependence plots to look into the dependence of these features on the continuous variables, i.e., age and the interval between the two doses (**Fig 2B**). All the features decreased as age increased. Two features, the AUC and the peak, increased as the interval increased. By contrast, the duration was smallest at the interval of 21 days and increased as the interval became longer than that. However, we had only 13 participants with an interval of fewer than 20 days and the difference between their duration and the duration of others was not statistically significant (*p* = 0.61). We next looked at the dependence of the features on categorical variables: we found that the presence of underlying diseases (collagen diseases and rheumatism) and medication use (antihistamines, immunosuppressants, and steroids) affected the features negatively, while the presence of adverse reactions affected the features positively. The result of a similar analysis on the model parameters *H*_2_ and *m* is shown in **Supplementary Fig 2B**.

### Deriving a personalized antibody score

Combining the above demographic and health information, we devised a simple score that enables individuals to roughly estimate their antibody status. We chose the AUC of the antibody dynamics as a representative feature of individual antibody status, because the other features (the peak and the duration) are highly correlated with and similar to the AUC (**Fig 1C**). Recently, a systematic approach to fit optimized scores with mixed-integer nonlinear programming was proposed (19). We applied this method to construct two types of scores to cover the whole range of AUC: a score to predict whether an individual’s AUC is in the top third of the population (i.e., top AUC score, **Fig 2C**) and a score to predict whether an individual’s AUC is in the bottom third (i.e., bottom AUC score, **Supplementary Fig 2C**).

We first divided our dataset into training and test datasets. The training dataset consisted of participants in Minami Soma City and Hirata Village (1775 individuals), and the test dataset consisted of participants in Soma City (384 individuals). We used the training dataset for fitting the scores. The algorithm searches the space of linear combinations of features with integer coefficients from -5 to +5 to find the best combination to differentiate the population in the top third of the training dataset from the rest (or the bottom third from the rest) (**Fig 2C**). We then assessed the performance of these scores in the test dataset, that is, we tested whether the scores just created could differentiate the population in the top third of the test dataset from the rest (or the bottom third from the rest).

The top AUC scores in the test dataset were between -2 and 2, except for two individuals with scores of -5 and 4, respectively (**Fig 2D** left). The 91 individuals with scores of 2 or more (shown in yellow and orange) included 52 individuals (shown in yellow) whose AUC belonged to the top third of the test dataset population (57.1%). The 104 individuals with a score of 1 included 46 individuals with AUC in the top third (44.2%). The 92 individuals with a score of 0 included 13 individuals with AUC in the top third (14.1%). The 78 individuals with a score of -1 included 16 individuals with AUC in the top third (20.5%). The 19 individuals with a score of -2 or less included only 1 individual with AUC in the top third (5.2%). Thus, the higher an individual’s top AUC score, the more likely they were to belong to the top third of the population and have a higher AUC as well (**Fig 2D** right). On the other hand, we calculated the bottom AUC scores in the test dataset, which were between -4 and 2, except for one individual with a score of 3 (**Supplementary Fig 2D** left). We confirmed similar trends. For example, the 65 individuals with a score of 1 included 34 individuals with AUC in the bottom third (52.3%). Thus, the higher an individual’s bottom AUC score, the more likely they were to belong to the bottom third of the population and have a lower AUC as well (**Supplementary Fig 2D** right). These results suggest that the personalized AUC score can estimate individual antibody status with reasonable accuracy, helping individuals make informed decisions about their disease prevention.

## Discussion

In this study, we created a personalized antibody score that evaluates the antibody status of individuals. To make an optimal score, we used a mathematical model of antibody production in response to two-dose mRNA vaccinations, as developed in our previous paper (18), and reconstructed the vaccine-elicited antibody dynamics of 2,159 participants from the Fukushima vaccination cohort. Our mechanism-based mathematical modeling, in contrast to the statistical modeling used in recent reports (3, 20), enabled biologically accurate description and precise comparison of antibody dynamics. The parameters of the estimated dynamics showed a large variation spanning two orders of magnitude. This variation was partially explained by individual characteristics like age, sex, the interval between the two vaccine doses, adverse reactions, comorbidities, and medications taken. This result is consistent with previous studies reporting age, sex, vaccine interval, and comorbidities as factors affecting antibody titers (21, 22). Quantifying the variability in antibody dynamics can be a basis for policy decisions regarding the distribution of booster vaccines to strengthen immunity (23) or the use of oral antiviral drugs for the treatment of breakthrough infections (24).

Our antibody scores can be easily calculated from individual demographic and health information, yet have the ability to identify participants with high and low antibody titers (AUC of the IgG(S) titers). Given the pleiotropic aspect of humoral immunity, it is surprising that a score consisting of only 10 questionnaire items can give reasonable predictions. The score showed that COVID-19 infection history and adverse reactions positively affect the AUC, whereas age, diabetes, and collagen diseases negatively affect the AUC. These positive and negative factors are consistent with previous studies (22, 25-28). In addition, diabetes and autoimmune diseases have been reported to be risk factors for breakthrough infections (29, 30). Considering that individual antibody titers are partially predictive of the likelihood of breakthrough infections (31, 32), this suggests that our antibody score could also be used as a risk score for breakthrough infections. On the other hand, the antibody score also has some similarities with COVID-19 severity scores (33-37). In fact, both include age as a factor likely to lead to low antibody titers and critical illness, both of which may be related to a defective immune system, as observable in cytokine signatures (38, 39) or immunoglobulin responses (40, 41). However, whereas COVID-19 severity scores use the results of laboratory tests and clinical symptoms to assess the patient’s condition in the hospital, our antibody score can be calculated on the basis of questionnaire responses provided by individual (i.e., not limited to patients) themselves.

There are limitations to this study. The model fitting was based on limited antibody measurements (two or three times for most participants) and the antibody score was solely based on information available from the questionnaire. Further refinement of the score using additional information will be a worthwhile task. It is worth mentioning that the immune system is affected by multiple factors, including genetics, the environment (such as cohabitation), and markers of metabolic health (42-45), all of which likely influence individual antibody status but were not considered here. The biological determinants of antibody variation will be further revealed in future studies addressing not only B cell subsets but the whole immune system encompassing adaptive as well as innate immunity. At this stage, our score would best be used by medical practitioners as a tool to advise individuals on getting booster vaccinations or taking additional precautions against infection.

## Methods

### Study data

This study was conducted from April 2021 to December 2021 in Fukushima, Japan (called the Fukushima vaccination cohort). A total of 2,526 participants who had been vaccinated with Pfizer BNT162b2 or Moderna mRNA-1273 were recruited, and 2,159 participants were included in the final data analysis. The participants included health care workers, frontline workers, administrative officers, general residents, and residents of long-term care facilities. We here investigated antibody titers of individuals sampled longitudinally (serum was collected at 2 or 3 different timepoints) for around 4 to 9 months after the second primary dose of mRNA vaccine. Information on sex, age, daily medication, medical history, date of vaccination, adverse reactions after vaccination, type of vaccination, blood type, Bacillus Calmette–Guérin (BCG) vaccine history, smoking habits, and drinking habits were retrieved from the paper-based questionnaire (summarized in **Supplementary Table 1**). In addition to the participants in the cohort, we included 12 health care workers whose serum had been sequentially sampled for 40 days (on average 25 samples per individual) in our analysis for the validation and parameterization of a mathematical model for vaccine-elicited antibody dynamics. All serological assays were conducted at The University of Tokyo. (S)-specific IgG (i.e., IgG(S)) and neutralizing activity were measured as the humoral immune status after the COVID-19 vaccination. (N)-specific IgG antibody titers (IgG(N)) were used to determine past COVID-19 infection status (46). The study was approved by the ethics committees of Hirata Central Hospital (number 2021-0611-1) and Fukushima Medical University School of Medicine (number 2021-116). Written informed consent was obtained from all participants individually before the survey. A portion of this cohort was described previously for the period extending to 6 months after the first dose of mRNA vaccine (46, 47).

### Modeling vaccine-elicited antibody dynamics

We recently developed a mathematical model describing COVID-19 vaccine-elicited antibody dynamics to evaluate the impact of primary two-dose COVID-19 vaccination on rapid immunity at the individual level and reconstructed the best-fit antibody titer curves of 2,407 participants in the Fukushima cohort (18). Here we explain the derivation and formulation of the mathematical model in detail.

### (i) Vaccination-elicited antibody dynamics after the first dose

After the first vaccination, naïve B cells encounter the antigens and differentiate into short-lived antibody-secreting cells (ASCs), plasmablasts, germinal center (GC) B cells, or GC-independent memory B cells depending on BCR affinity for their cognate antigen (48). Then, the GC B cells undergo rapid proliferation with somatic immunoglobulin hypermutation and subsequently differentiate into GC-dependent memory B cells or long-lived antibody-secreting cells, which are plasma cells with immunoglobulin class switching. To describe this antigen-specific B cell expansion and the induction of antibody-secreting cells and memory B cells after the first vaccination (**Supplementary Fig 3A**), we developed a simple but quantitative mathematical model as follows:

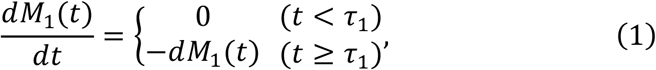

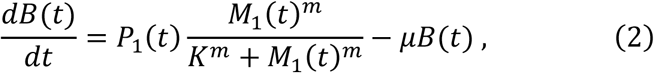

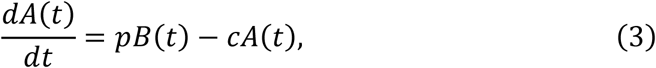

where the variables *M*_1_(*t*), *B*(*t*), and *A*(*t*) are the amount of mRNA inoculated by the vaccination, the number of antibody-secreting cells, and the antibody titers at time *t*, respectively. The parameters *τ*_1_ and *d* represent the timing of the vaccination and the decay rate of mRNA. We here considered *D*_1_ to be the inoculated dose of mRNA by the vaccination, that is, *M*_1_(*τ*_1_) = *D*_1_.

Because the data we used here were limited (i.e., only time-course vaccine-elicited IgG(S) titers), one compartment of B cells including heterogeneous cell populations that produce antibodies (i.e., short-lived and long-lived antibody-secreting cells) was assumed. Therefore, we modelled the average B cell population dynamics in Eq.(2), where the product of *P*_1_(*t*) and *M*_1_(*t*)^*m*^/(*M*_1_(*t*)^*m*^ + *K*^*m*^) represents the average *de novo* induction of the antibody-secreting cells. *P*_1_(*t*) is a step function defined as *P*_1_(*t*) = *P*_1_ for *τ*_1_ + *η*_1_ ≤ *t*, where *η*_1_ is the delay of induction of antibody-secreting cells after vaccination: otherwise *P*_1_(*t*) = 0. The parameters *m, K*, and *μ* correspond to the steepness at which the induction increases with increasing amount of mRNA (i.e., the Hill coefficient), the amount of mRNA satisfying *P*_1_/2, and the average decay rate of the antibody-secreting cell compartment, respectively. The other parameters, *p* and *c*, represent the antibody production rate and the clearance rate of antibodies, respectively.

### (ii) Vaccination-elicited antibody dynamics after the second dose

After the second vaccination, the memory B cells are reactivated by re-exposure to the antigen. Some differentiate into short-lived antibody-secreting cells (plasmablasts) or memory B cells outside the GC. Others enter the GC to become secondary GC B cells. Subsequently, these secondary GC B cells differentiate into GC-dependent memory B cells or long-lived antibody-secreting cells (plasma cells). To describe these recall B cell responses and their secretion of antibody after the second vaccination (**Supplementary Fig 3B**), we modified the above mathematical model, Eqs.(1-3), as follows:

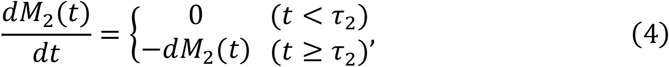

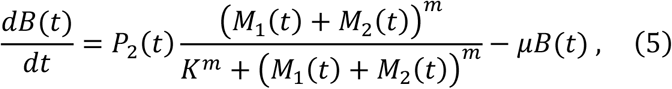

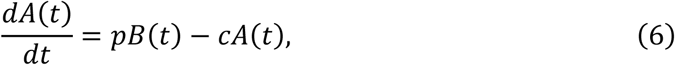

where *M*_2_(*t*) is the amount of mRNA by the vaccination inoculated at *τ*_2_ satisfying *M*_2_(*τ*_2_) = *D*_2_. In addition, *P*_2_(*t*) = *P*_1_ for *τ*_2_ ≤ *t* < *τ*_2_ + *η*_2_, *P*_2_(*t*) = *P*_2_ for *τ*_2_ + *η*_2_ ≤ *t*, where *η*_2_ is the delay of induction of antibody-secreting cells after vaccination; otherwise *P*_2_(*t*) = 0. It should be noted that, prior to this main recall immunity, some rapid but small reactivation is observed, probably due to GC-independent memory B cells induced by the first vaccination (see participants S2, S7, and S8 in **Supplementary Fig 1A**). This small reactivation is described by *P*_1_(*M*_1_(*t*) + *M*_2_(*t*))^*m*^/ ((*M*_1_(*t*) + *M*_2_(*t*))^*m*^ + *K*^*m*^) for *τ*_2_ ≤ *t* < *τ*_2_ + *η*_2_. In general, once reactivated, memory B cells can reenter the GC more rapidly than naïve B cells, and therefore the secondary antibody responses are much faster and larger (i.e., *η*_1_ > *η*_2_ and *P*_1_ < *P*_2_, respectively). In the main recall immunity, the quantity and quality of memory B cells established by the first vaccination is included in *P*_2_.

### (iii) Mathematical model for data fitting

Since the clearance rate of antibody is much larger than the decay of antibody-secreting cells (i.e., *c* ≫ *μ*), we made a quasi-steady state assumption, *dA*(*t*)/*dt* = 0, and replaced Eqs.(3) and (6) with *A*(*t*) = *pd*(*t*)/*c*. Moreover, since Eqs.(1) and (4) are the linear differential equations, *M*_1_(*t*) = *D*_1_*e*^−*dt*^ for *t* ≥ *τ*_1_ and *M*_2_(*t*) = *D*_2_*e*^−*dt*^ for *t* ≥ *τ*_2_ : otherwise *M*_1_(*t*) = *M*_2_(*t*) = 0, respectively. Thus, the above Eqs.(1-6) are further simplified assuming *τ*_1_ = 0 and *D*_1_ > 0, and we obtained the following single ordinary differential equation, which we used to analyze the antibody responses (i.e., IgG(S) titers (AU/mL)) in this study:

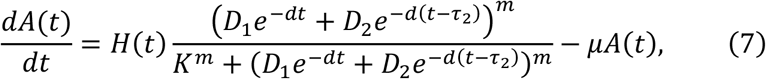

where *H*(*t*) = *H*_*i*_ = *pP*_*i*_/*c* for *τ*_*i*_ + *η*_*i*_ ≤ *t* < *τ*_*i*+1_ + *η*_*i*+1_ (*i* = 1 or 2) and *D*_2_ > 0 for *τ*_2_ < *t* : otherwise *D*_2_ = 0. This simple model can quantify the vaccine-elicited time-course antibody dynamics as described in **Fig 1A** under an arbitrary threshold of antibody titers *A*_TH_ (see below).

### Parameter estimations

To evaluate the primary two-dose COVID-19 vaccination leading to the antibody titers (i.e., IgG(S) titers), we used a nonlinear mixed effects model to fit the antibody dynamics model, given by Eq.(7), to the longitudinal antibody titers of IgG(S) obtained from the 12 health care workers. Briefly, the parameters for individual *k*, 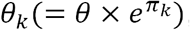, are represented as a product of *θ* (a fixed effect) and 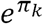 (a random effect). *π*_*k*_ follows a normal distribution with mean 0 and standard deviation Ω. We here assumed that the parameters *H*_1_, *H*_2_, *η*_1_, *η*_2_, and *m* vary across individuals, although we do not consider the inter-individual variability in other parameters to ensure parameter identifiability. Note that the half-life of mRNA (i.e., log 2 /*d*) and dose of mRNA (i.e., *D*_*i*_) are assumed to be 1 day (49) and 100 (*μμ*/0.5mL) (50), respectively. We estimated fixed effects and random effects using the stochastic approximation expectation-approximation algorithm and empirical Bayes’ method, respectively. Fitting was performed using MONOLIX 2019R2 (www.lixoft.com) (51). The estimated (fixed and individual) parameters are listed in **Supplementary Table 2**.

With the estimated parameters for each individual, the dynamics of IgG(S) titers, *A*(*t*), and the average *de novo* antibody response elicited by the first and second vaccinations, 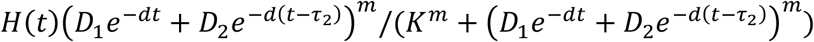, were calculated in **Supplementary Fig 1A** and **1B**, respectively. Interestingly, we observed that the variations induced by the second vaccination were much larger than those induced by the first vaccination (**Supplementary Fig 1B**). Although we found that most of the best-fitted estimated parameters in the mathematical model (i.e., *μ, K, η*_1_, *η*_2_, *H*_1_) were the same or similar across the 12 individuals, the parameters *m* and *H*_2_, which contribute mainly to the vaccine-elicited antibody dynamics after the second vaccination, showed wide variation of estimated values (see **Supplementary Table 2**). Therefore, fixing these estimated population parameters except for *m* and *H*_2_, we applied a nonlinear least squares method to reconstruct the large variations of the antibody dynamics after the second dose for 2,159 participants. The best-fit antibody titer curves are plotted along with the observed data for visualization in **Supplementary Fig 1C**, and the distribution of parameter values *m* and *H*_2_ are summarized in **Supplementary Fig 1D**.

### Random forest regressors for characterizing vaccine-elicited antibody dynamics

Random forest regressors were trained to predict any of the three features of antibody dynamics (log of peak, log of AUC, duration) on the basis of the participants’ demographic and health information as obtained by questionnaire (see Statistical analysis). randomForest and rfPermute packages in R were used. The R squared values for each regressor were calculated from OOB samples. Feature importance, which shows to what extent each factor was predictive of the antibody dynamics features, was based on percentage increase in mean squared errors and is shown as Chord diagrams (circlize package in R), in which arrows are drawn from each feature to its predictive factors in a circular layout. Only the factors with *p* < 0.05 (obtained with 1,000 permutations) were selected and shown.

### Building optimized antibody scores

A Python implementation of Ustun et al.’s (19) algorithm (risk-slim, https://github.com/ustunb/risk-slim) was used to build optimized AUC scores. Briefly, the algorithm searches for the best linear combination of features with integer coefficients that minimizes the sum of the logistic loss and the *l*_0_-norm of the coefficients. The range of coefficients was set to -5 to +5; the *l*_0_-penalty parameter C_0_ was set to 5×10^−4^.

### Statistical analysis

Answers to the paper-based questionnaire collected from 2,159 participants were converted into a set of categorical and numerical variables. Numerical variables included age and the interval between the two doses. These variables were then used as input to predict the three antibody dynamics features. Missing values of categorical variables were treated as a separate category and were included in the analyses. The variables used here belonged to any of the five categories: (i) basic demographic information and lifestyle habits, (ii) information on vaccinations, (iii) underlying medical conditions, (iv) adverse reactions, and (v) medications being taken. When necessary, the same variables were compared among different generations or different groups using Pearson’s chi-square test (for categorical variables), analysis of variance (ANOVA, for more than two numerical variables), or Welch T-test (for two numerical variables). Pearson correlation coefficient was calculated to evaluate the association between a pair of continuous variables. All statistical analyses were performed using R (version 4.1.2).

## Data Availability

All data produced in the present study are available upon reasonable request to the authors

## List of supplementary materials

**Supplementary Figure 1.**
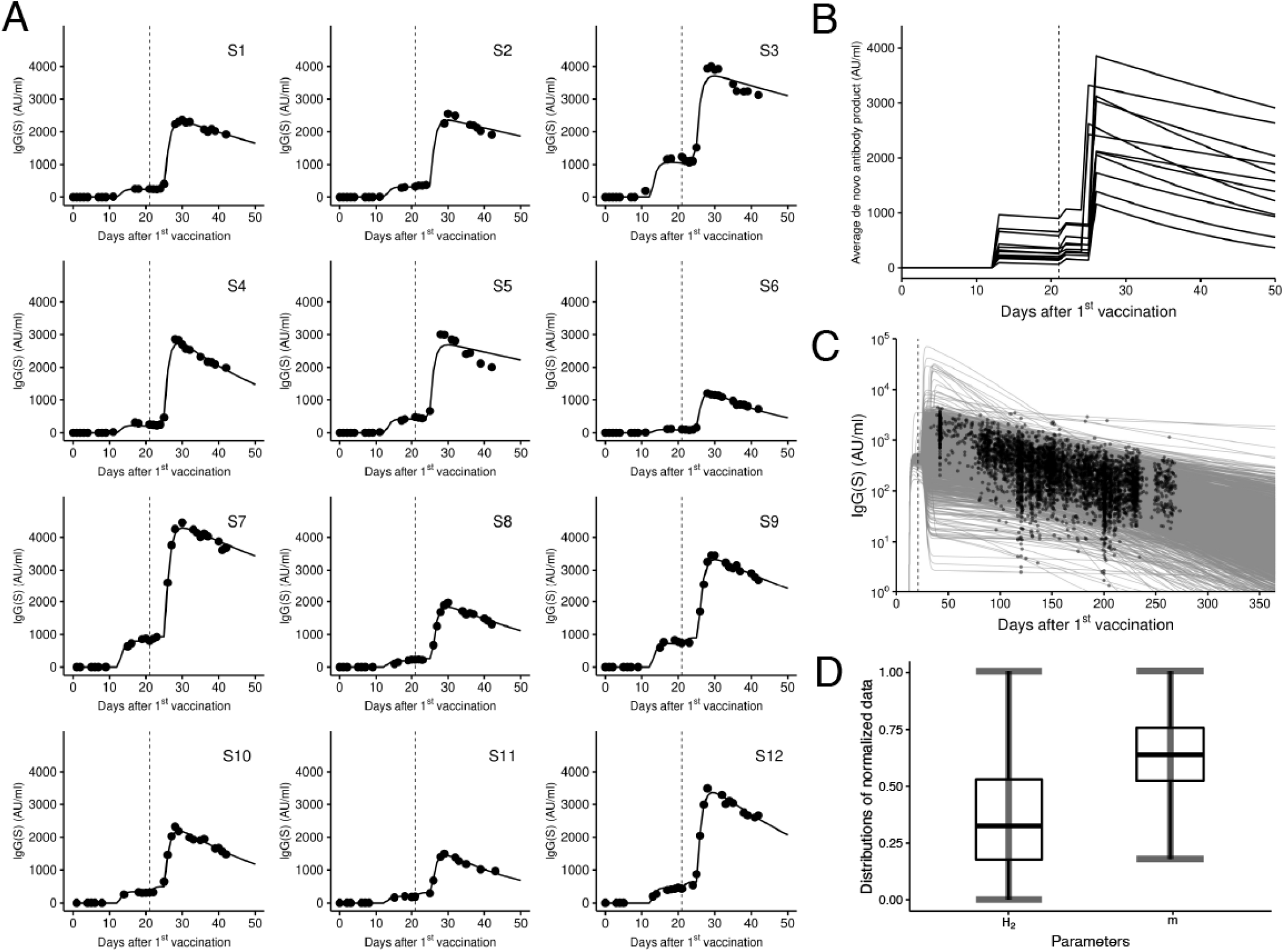
Calibrating vaccine-elicited antibody dynamics: **(A)** Observed and best-fitted IgG(S) titers are described for the 12 health care workers (HCWs) whose serum was sequentially sampled. The dashed vertical lines at day 21 correspond to the date of second vaccination. **(B)** Time-course averages of *de novo* antibody response elicited by the first and second vaccinations for the 12 HCWs are described. **(C)** Reconstructed individual antibody dynamics for the 2,159 participants are represented along with the measured IgG(S). The black circles correspond to the measurements of antibody titers at different time points. **(D)** Distributions of the estimated parameter values (i.e., *H*_2_ and *m*) for 2,407 participants are plotted. Dataset for each distribution is normalized by the value corresponding to the 95th percentile of data values, and data whose values were larger than this value were removed to improve visibility of the figure.

**Supplementary Figure 2.**
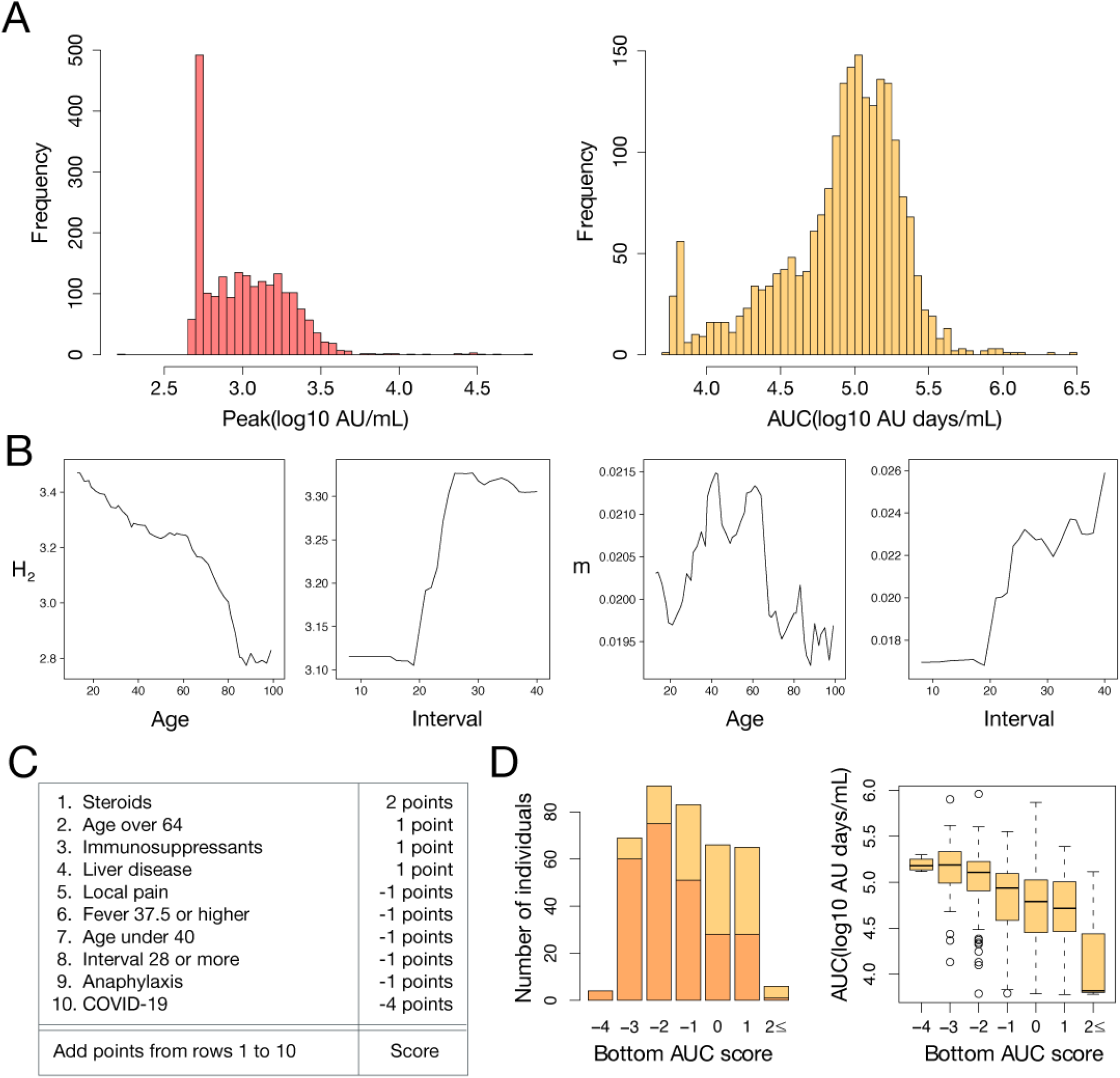
Analyzing antibody titers: **(A)** The distributions of two features of antibody dynamics (peak and AUC) in the cohort are shown. **(B)** Partial dependence plots showing the dependence of the two parameters (*H*_2_ and *m*) on age or the interval are shown. **(C)** The bottom AUC score to identify individuals with AUC in the bottom third of the population is shown. **(D)** Left: The distribution of the bottom AUC score in the test dataset. 4, 69, 91, 83, 66, 65, and 6 individuals had scores of -4, -3, -2, -1, 0, 1, or 2 or more, respectively. Those in the bottom third of the test dataset are shown in yellow, and those not in the bottom third are shown in orange. The ratio of individuals with AUC in the bottom third of the test dataset increased as the bottom AUC score increased. Right: the average AUC tended to decrease as the bottom AUC score increased.

**Supplementary Figure 3.**
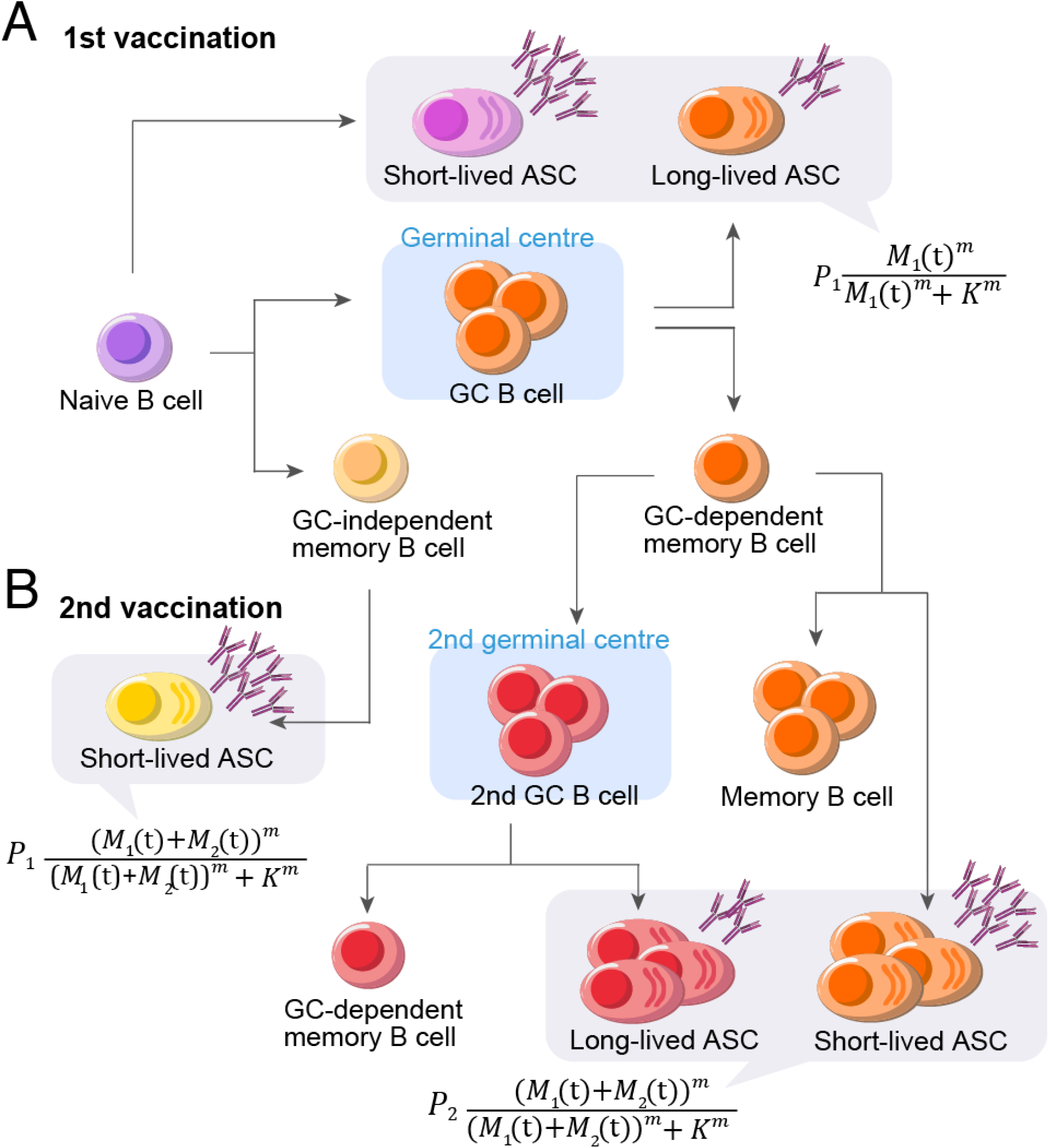
Modeling vaccine-elicited B cell dynamics: **(A)** First vaccination-elicited antibody-secreting cell and memory B cell inductions are described. Once naïve B cells encounter vaccine antigens outside the germinal center (GC), the activated naïve B cells differentiate into short-lived antibody-secreting cells (ASCs), plasmablasts, GC B cells, or GC-independent memory B cells depending on BCR affinity for their cognate antigen. Subsequently, the GC B cells undergo rapid proliferation with somatic immunoglobulin hypermutation and differentiate into GC-dependent memory B cells or long-lived antibody-secreting cells (plasma cells), with immunoglobulin class switching. Through the GC-independent and dependent pathways, antibody-secreting cells (i.e., *B*(*t*)) are induced and they secrete antibodies (i.e., *A*(*t*)).**(B)** Second vaccination-elicited recall immune responses are described. After re-exposure to vaccine antigens, memory B cells rapidly reactivate and expand. Of activated memory B cells, while some differentiate into plasmablasts or memory B cells outside the GC, others enter the GC to be secondary GC B cells. These secondary GC B cells differentiate into GC-dependent memory B cells or plasma cells. In general, the secondary antibody responses are much faster and larger by an order of magnitude compared with the first vaccination-elicited antibody responses.

**Supplementary Table 1.** Basic demographics for the Fukushima vaccination cohort.

| Characteristic | <40 years | 40-64 years | >65 years | Overall | p-value |
| --- | --- | --- | --- | --- | --- |
| **Gender** |  |  |  |  | 0.001 |
| Male | 338(46.8) | 421(38.2) | 286(40.8) | 1045(41.4) |  |
| **Vaccine type** |  |  |  |  | 0.015 |
| BNT162b2 (Pfizer–BioNTech) | 651(90.0) | 971(88.1) | 649(92.6) | 2271(89.9) |  |
| mRNA-1273 (Moderna) | 0(0.0) | 3(0.3) | 0(0.0) | 3(0.1) |  |
| **Days (mean [SD])** |  |  |  |  |  |
| from 1st dose | 106.3[37.7] | 104.4[35.1] | 103.8[17.2] | 104.8[32.0] | 0.302 |
| from 2nd dose | 181.4[37.5] | 180.1[35.7] | 180.1[16.9] | 180.5[32.2] | 0.292 |
| **Blood type** |  |  |  |  |  |
| A | 239(35.7) | 427(39.0) | 212(35.4) | 878(37.2) | <0.001 |
| B | 151(22.6) | 236(21.6) | 140(23.4) | 527(22.3) | <0.001 |
| O | 213(31.8) | 326(29.8) | 172(28.7) | 711(30.1) | 0.367 |
| AB | 66(9.9) | 105(9.6) | 75(12.5) | 246(10.4) | 0.432 |
| **BCG history** | 560(83.5) | 907(83.1) | 412(61.9) | 1879(77.4) | 0.395 |
| **Smoking** | 144(19.9) | 253(23.0) | 63(9.0) | 460(18.2) | <0.001 |
| **Drinking Habit** |  |  |  |  | <0.001 |
| Almost not | 403(55.7) | 542(49.2) | 458(65.3) | 1403(55.5) |  |
| Occasionally | 247(34.2) | 310(28.1) | 90(12.8) | 647(25.6) |  |
| Everyday | 63(8.7) | 222(20.2) | 127(18.1) | 412(16.3) |  |
| **Daily Alcohol Consumption** |  |  |  |  | <0.001 |
| <20g | 322(44.5) | 382(34.7) | 176(25.1) | 880(34.8) |  |
| 20-40g | 108(14.9) | 214(19.4) | 87(12.4) | 409(16.2) |  |
| 40-60g | 20(2.8) | 60(5.4) | 20(2.9) | 100(4.0) |  |
| >60g | 5(0.7) | 13(1.2) | 2(0.3) | 20(0.8) |  |
| **Comorbidities** |  |  |  |  |  |
| Hypertension | 8(1.1) | 237(21.5) | 432(61.6) | 677(26.8) | <0.001 |
| Dyslipidemia | 12(1.7) | 123(11.2) | 146(20.8) | 281(11.1) | <0.001 |
| Heart disease | 14(1.9) | 47(4.3) | 140(20.0) | 201(8.0) | <0.001 |
| Diabetes | 6(0.8) | 72(6.5) | 110(15.7) | 188(7.4) | <0.001 |
| Allergic disease | 69(9.5) | 95(8.6) | 21(3.0) | 185(7.3) | <0.001 |
| Asthma | 49(6.8) | 45(4.1) | 28(4.0) | 122(4.8) | 0.079 |
| Liver disease | 11(1.5) | 45(4.1) | 58(8.3) | 114(4.5) | <0.001 |
| Cancer | 3(0.4) | 35(3.2) | 46(6.6) | 84(3.3) | <0.001 |
| Gout | 5(0.7) | 45(4.1) | 26(3.7) | 76(3.0) | 0.001 |
| Thyroid disease | 8(1.1) | 40(3.6) | 11(1.6) | 59(2.3) | 0.005 |
| Lung disease | 12(1.7) | 11(1.0) | 28(4.0) | 51(2.0) | <0.001 |
| Mental disease | 17(2.4) | 16(1.5) | 13(1.9) | 46(1.8) | 0.739 |
| Rheumatism | 2(0.3) | 16(1.5) | 19(2.7) | 37(1.5) | 0.006 |
| Kidney disease | 6(0.8) | 7(0.6) | 14(2.0) | 27(1.1) | 0.089 |
| Anaphylaxis | 6(0.8) | 7(0.6) | 5(0.7) | 18(0.7) | 0.994 |
| Collagen disease | 4(0.6) | 6(0.5) | 5(0.7) | 15(0.6) | 0.993 |
| COVID-19 (family) | 4(0.6) | 5(0.5) | 1(0.1) | 10(0.4) | 0.793 |
| COVID-19 | 0(0.0) | 3(0.3) | 4(0.6) | 7(0.3) | 0.38 |
| Immune deficiency | 2(0.3) | 4(0.4) | 0(0.0) | 6(0.2) | 0.654 |
| Others | 51(7.1) | 147(13.3) | 189(27.0) | 387(15.3) | <0.001 |
| **Drug** |  |  |  |  |  |
| Steroid | 9(1.2) | 23(2.1) | 26(3.7) | 58(2.3) | 0.002 |
| NSAIDs | 31(4.3) | 78(7.1) | 82(11.7) | 191(7.6) | <0.001 |
| Acetaminophen | 8(1.1) | 22(2.0) | 30(4.3) | 60(2.4) | <0.001 |
| Antihistamine | 46(6.4) | 65(5.9) | 43(6.1) | 154(6.1) | 0.367 |
| Immunosuppressants | 6(0.8) | 10(0.9) | 8(1.1) | 24(1.0) | 0.432 |
| Biologics | 2(0.3) | 5(0.5) | 4(0.6) | 11(0.4) | 0.395 |
| Anti-cancer agent | 0(0.0) | 5(0.5) | 5(0.7) | 10(0.4) | 0.292 |

| **Adverse Reaction** |  |  |  |  |  |
| --- | --- | --- | --- | --- | --- |
| Local pain | 515(71.2) | 684(62.1) | 228(32.5) | 1427(56.5) | <0.001 |
| Fatigue | 511(70.7) | 627(56.9) | 119(17.0) | 1257(49.8) | <0.001 |
| Joint pain | 327(45.2) | 354(32.1) | 90(2.8) | 771(30.5) | <0.001 |
| Fever (37.5 degrees or higher) | 370(51.2) | 308(28.0) | 41(5.9) | 719(28.5) | <0.001 |
| Headache | 321(44.4) | 331(30.0) | 34(4.9) | 686(27.2) | <0.001 |
| Fever (under 37.5 degrees) | 137(19.0) | 209(19.0) | 40(5.7) | 386(15.3) | <0.001 |
| Dizziness | 57(7.9) | 45(4.1) | 9(1.3) | 111(4.4) | <0.001 |
| Nausea | 51(7.1) | 41(3.7) | 6(0.9) | 98(3.9) | <0.001 |
| Diarrhea | 30(4.2) | 25(2.3) | 3(0.4) | 58(2.3) | <0.001 |
| Others | 40(5.5) | 69(6.3) | 16(2.3) | 125(5.0) | 0.003 |

Values are No. (%) unless noted otherwise. BCG, bacille Calmette-Guérin; NSAID, nonsteroidal anti-inflammatory drug.

**Supplementary Table 2.**
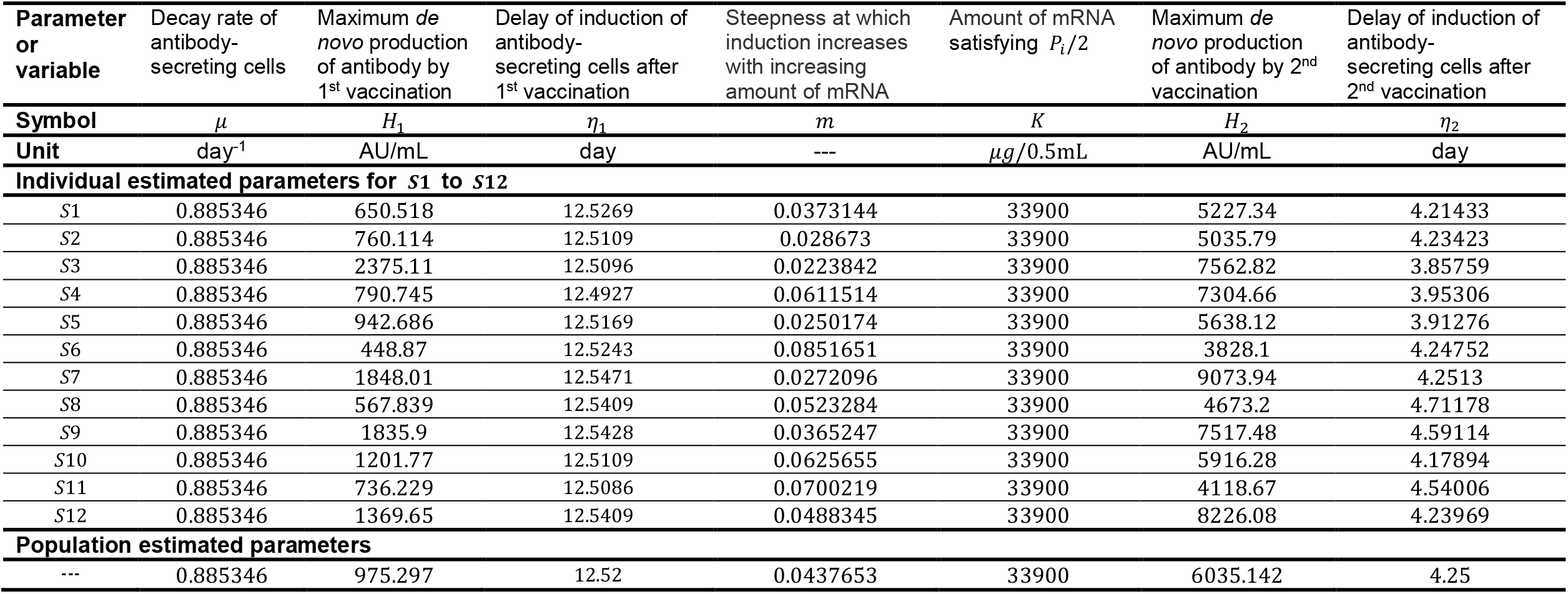
Estimated fixed and individual parameters for 12 health care workers.

| Parameter or variable | Decay rate of antibody-secreting cells | Maximum *de novo* production of antibody by 1<sup>st</sup> vaccination | Delay of induction of antibody-secreting cells after 1<sup>st</sup> vaccination | Steepness at which induction increases with increasing amount of mRNA | Amount of mRNA satisfying $P_i/2$ | Maximum *de novo* production of antibody by 2<sup>nd</sup> vaccination | Delay of induction of antibody-secreting cells after 2<sup>nd</sup> vaccination |
| --- | --- | --- | --- | --- | --- | --- | --- |
| Symbol | $\mu$ | $H_1$ | $\eta_1$ | $m$ | $K$ | $H_2$ | $\eta_2$ |
| Unit | day<sup>-1</sup> | AU/mL | day | --- | $\mu\text{g}/0.5\text{mL}$ | AU/mL | day |
| **Individual estimated parameters for $S_1$ to $S_{12}$** |  |  |  |  |  |  |  |
| $S_1$ | 0.885346 | 650.518 | 12.5269 | 0.0373144 | 33900 | 5227.34 | 4.21433 |
| $S_2$ | 0.885346 | 760.114 | 12.5109 | 0.028673 | 33900 | 5035.79 | 4.23423 |
| $S_3$ | 0.885346 | 2375.11 | 12.5096 | 0.0223842 | 33900 | 7562.82 | 3.85759 |
| $S_4$ | 0.885346 | 790.745 | 12.4927 | 0.0611514 | 33900 | 7304.66 | 3.95306 |
| $S_5$ | 0.885346 | 942.686 | 12.5169 | 0.0250174 | 33900 | 5638.12 | 3.91276 |
| $S_6$ | 0.885346 | 448.87 | 12.5243 | 0.0851651 | 33900 | 3828.1 | 4.24752 |
| $S_7$ | 0.885346 | 1848.01 | 12.5471 | 0.0272096 | 33900 | 9073.94 | 4.2513 |
| $S_8$ | 0.885346 | 567.839 | 12.5409 | 0.0523284 | 33900 | 4673.2 | 4.71178 |
| $S_9$ | 0.885346 | 1835.9 | 12.5428 | 0.0365247 | 33900 | 7517.48 | 4.59114 |
| $S_{10}$ | 0.885346 | 1201.77 | 12.5109 | 0.0625655 | 33900 | 5916.28 | 4.17894 |
| $S_{11}$ | 0.885346 | 736.229 | 12.5086 | 0.0700219 | 33900 | 4118.67 | 4.54006 |
| $S_{12}$ | 0.885346 | 1369.65 | 12.5409 | 0.0488345 | 33900 | 8226.08 | 4.23969 |
| **Population estimated parameters** |  |  |  |  |  |  |  |
| --- | 0.885346 | 975.297 | 12.52 | 0.0437653 | 33900 | 6035.142 | 4.25 |

## Acknowledgments

We would like to thank all the staff from Fukushima Medical University, Seireikai health care group, Hirata Village office, Soma City office, Soma Central Hospital, Soma General Hospital, Minamisoma City office, Minamisoma City Medical Association, Minamisoma Municipal General Hospital, Shindo Clinic and Medical Governance Institute, who contributed significantly to the accomplishment of this research, especially, Ms. Yuka Harada, Ms. Serina Noji, Ms. Naomi Ito, Dr. Makoto Kosaka, Mr. Anju Murayama, Mr. Sota Sugiura, Mr. Manato Tanaka, Ms. Yuna Uchi, Mr. Yudai Kaneda, Mr. Masahiko Nihei, Mr. Hideo Sato, Ms. Rie Yanai, Ms. Yasuko Suzuki, Ms. Keiko Abe, Dr. Hidekiyo Tachiya, Mr. Kouki Nakatsuka, Dr. Ryuzaburo Shineha, Ms. Miki Sato, Dr. Masahiko Sato, Mr. Naoharu Tadano, Mr. Kazuo Momma, Mr. Shu-ichi Mori, Ms. Saori Yoshisato, Ms. Katsuko Onoda, Mr. Satoshi Kowata, Mr. Masatsugu Tanaki, Dr. Tomoyoshi Oikawa, Dr. Joji Shindo, Ms. Xujin Zhu, Ms. Asaka Saito, Ms. Yuumi Kondo, and Ms. Tomoyo Nishimura. This study was supported by Medical & Biological Laboratories Co., Ltd., Shenzhen YHLO Biotech Co., Ltd., the distributor and manufacturer of the antibody measurement system (iFlash 3000), Research Center for Advanced Science and Technology in the University of Tokyo, and in part by Scientific Research (KAKENHI) B 18H01139 (to S.I.), 16H04845 (to S.I.); Scientific Research in Innovative Areas 20H05042 (to S.I.); AMED Strategic International Brain Science Research Promotion Program 22wm0425011s0302 (to K.A.); AMED JP22dm0307009 (to K.A.); AMED CREST 19gm1310002 (to S.I.); AMED Development of Vaccines for the Novel Coronavirus Disease, 21nf0101638h0001 (to M.T.), 21nf0101638s0201 (to S.I.); AMED Japan Program for Infectious Diseases Research and Infrastructure, 22wm0325007h0001 (to S.I.), 22wm0325004s0201 (to S.I.), 22wm0325012s0301 (to S.I.), 22wm0325015s0301 (to S.I.); AMED Research Program on Emerging and Re-emerging Infectious Diseases 22fk0108140s0802 (to S.I.); AMED Research Program on HIV/AIDS 22fk0410052s0401 (to S.I.); AMED Program for Basic and Clinical Research on Hepatitis 22fk0210094 (to S.I.); AMED Program on the Innovative Development and the Application of New Drugs for Hepatitis B 22fk0310504h0501 (to S.I.); AMED Research Program on Hepatitis 19fk0210036h0502 (to S.I.), 19fk0310114h0103 (to S.I.); JST MIRAI JPMJMI22G1 (to S.I.); Moonshot R&D JPMJMS2021 (to K.A. and S.I.) and JPMJMS2025 (to S.I.); The National Research Foundation of Korea (NRF) grant funded by the Korea government (MSIT) (2022R1C1C2003637) (to K.S.K.); Shin-Nihon of Advanced Medical Research (to S.I.); SECOM Science and Technology Foundation (to S.I.); The Japan Prize Foundation (to S.I.); and Kowa Co (to M.T.).

## Author contributions

SI designed the research. MT conducted the data collection. NN, HP, KSK, SIwanami, KA, and SI carried out the computational analysis. SI supervised the project. All authors contributed to writing the manuscript.

## Institutional review board statement

This study was approved by the ethics committees of Hirata Central Hospital (number 2021-0611-1) and Fukushima Medical University (number 2021-116). This study was conducted in accordance with the Code of Ethics of the World Medical Association (Declaration of Helsinki).

## Informed consent statement

Informed consent was obtained from all subjects involved in the study.

